# Surgeon Factors and Their Association With Operating Room Turnover Time

**DOI:** 10.1101/2024.09.10.24313267

**Authors:** Kshitij Pandit, Luke Wang, Joel Rosenberg, Nicole Goldhaber, Jill C. Buckley, Sonia Ramamoorthy, Kristin L. Mekeel, Aditya Bagrodia

## Abstract

**Introduction:** Operating room (OR) turnover time (TT), defined as the interval between the completion of one surgery and the start of the next, is a critical measure of OR efficiency impacting healthcare costs, patient outcomes, and surgical staff well-being. Previous research has identified various contributors to TT, such as surgical team dynamics, OR preparation, and interdisciplinary workflows. However, the influence of surgeon-specific factors like gender, administrative roles, and experience on TT remains underexplored. This study aims to address this gap by examining how these individual surgeon characteristics impact OR efficiency.

**Methods:** We conducted a retrospective study at the UC San Diego School of Medicine, a tertiary academic medical center. We analysed 12,820 surgical case entries from January 2022 to July 2023, sourced from the electronic health record system. Surgeons were categorized by gender, ethnicity, years of experience, training at UCSD, academic rank, and administrative roles. We utilized Mann-Whitney U test for binary variables and Kruskal-Wallis H test for variables with more than two categories. Multivariable linear regression was applied, adjusting for multiple comparisons using Holm correction. A p-value of less than 0.05 was considered statistically significant. All data analysis was performed using IBM SPSS version 29.

**Results:** Our analysis of 12,820 surgical cases revealed that surgeons in administrative roles and those with over ten years of experience demonstrated significantly shorter turnover times (TT). Specifically, administrators demonstrated a TT of 27 minutes, compared to 35 minutes for non-administrators (p<0.001) (Table 2). Surgeons with more than ten years of experience had a TT of 31 minutes, versus 37 minutes for those with less experience (p<0.001). Multivariable linear regression confirmed these associations, with significant reductions in TT linked to administrative roles (beta: - 7.2; 95% confidence interval (CI): -8.2 to -6.2, p<0.001) and surgeon experience (beta: -4.7, 95% CI: -5.9 to -3.5, p< 0.001).

**Conclusion:** We recommend efforts focusing on building a standardized environment for surgeons regardless of their background. This could lead to not only an equitable. OR culture but also an overall increase in the institution efficiency and patient outcomes.

## Introduction

Operating room (OR) Turnover time, the interval between the conclusion of one surgery and the start of the next is a key metric of an institution’s OR efficiency(1). Previous studies have explored multiple factors associated with turnover time, including surgical team composition, OR preparation, improved interdisciplinary work flow, surgeon involvement and surgical staff incentivization(2),(3),(4), thus identifying areas for targeted improvements in turnover time and enhancing OR output. However, the role of surgeon-specific factors on OR turnover time remains underexplored. Our study aims to fill this gap by examining the association of surgeon-specific factors with OR turnover time.

## Methods

We conducted a retrospective analysis of 12,820 surgical cases performed by 268 surgeons at a tertiary academic center. OR turnover time was calculated by summing six time intervals: 1) Wrap up time after end of first case 2) Wheeling out to clean-up start after the first case 3) Clean-up after first case 4) Completion of clean-up to the start of second case set-up 5) Second case set-up and, 6) Wheeling in patient for the second case. The variables captured were ethnicity, gender, experience (recorded as a binary variable as greater or less than 10 years of experience), residency or fellowship training at UCSD, academic rank and administrative position (Table 1). Administrative positions were defined if the surgeon served as the department chair or vice-chair, division chief or held a leadership role such as chief executive officer in the health administration system.

**Table 1:**
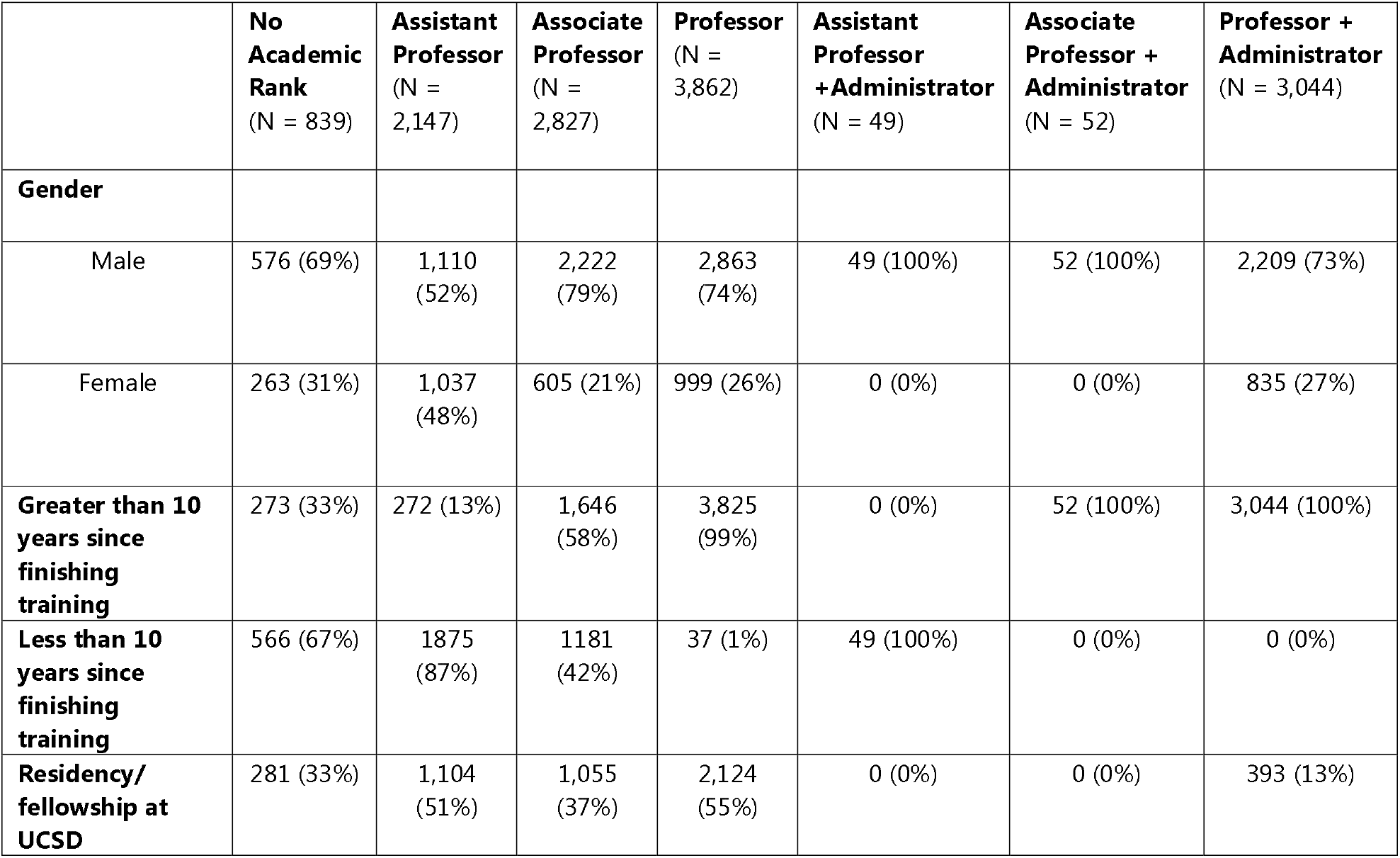
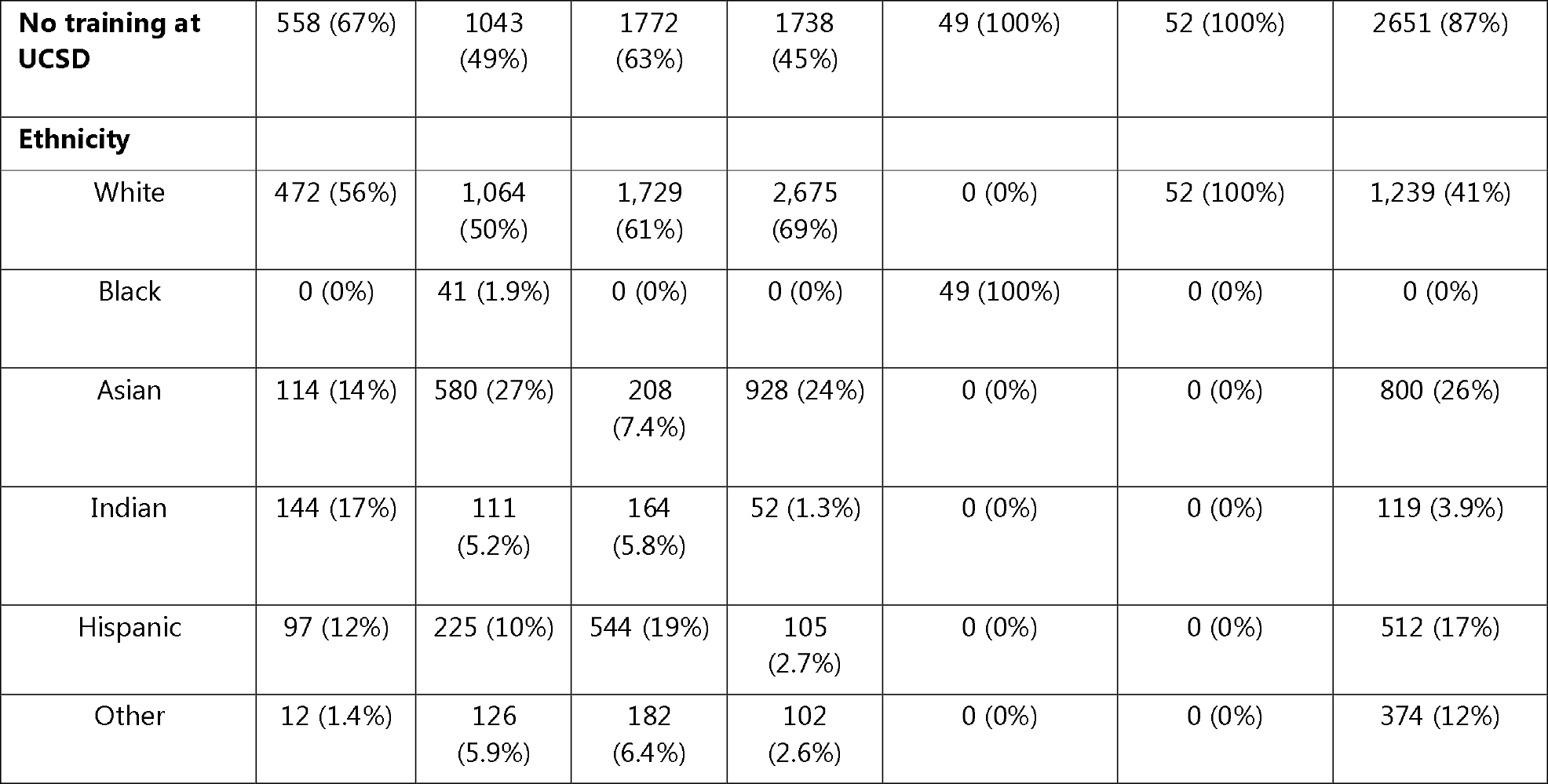
Surgeon demographics.

## Results

Surgeons with administrative roles were significantly associated with shorter turnover times compared to non-administrators (27 vs. 35 minutes, p<0.001). The turnover times for professors, associate and assistant professors with administrative roles were 27, 24.5 and 27 minutes, respectively. In comparison, surgeons without administrative positions demonstrated turnover times of 36, 35 and 38 minutes, respectively for the same academic ranks (p<0.001). Surgeons with ≥10 years of experience were also significantly associated with shorter turnover times (31 vs. 37 minutes, p<0.001). Multivariable linear regression confirmed these associations, with significant reductions in turnover time linked to administrative roles (β: -7.2; 95% confidence interval (CI): -8.2 to -6.2, p<0.001) and surgeon experience (β: -4.7, 95% CI: -5.9 to - 3.5, p< 0.001) (Table S1). Other factors such as gender, ethnicity and training of institution weren’t associated with significant differences in turnover times.

## Discussion

Our study distinguishes itself by providing a comprehensive analysis of surgeon-specific factors on OR turnover time. We would also like to encourage readers to consider our findings in the context of daily operations by highlighting that while a difference of 6-8 minutes in turnover time might seem trivial, it can accumulate over multiple cases, leading to significant time savings.

We hypothesize that multiple factors contribute to surgeons with leadership roles being associated with lower turnover times. Surgeons in administrative positions often have more control over policy implementation, choosing surgical teams and case scheduling, leading to more streamlined turnover practices. It was found that the presence of a fixed nurse team in the OR was associated with a shorter turnover times(5). The shorter turnover time amongst these surgeons could also be attributed to their understanding of logistics and better execution of these practices from the surgical team. While it has been established that surgical experience is positively corelated to improved surgical outcomes(6), there is sparse data on its impact on OR turnover time. Our finding of surgeons with greater than 10 years of experience being associated with shorter turnover time suggests that experience accumulated over the years is a contributing factor. Our findings highlighting non-significant associations of gender and ethnicity with turnover time are encouraging as they suggest equity within our institution.

Our study is not without its limitations. Due to the nature of data available, we could not control for the inherent differences that come with various procedure types and time of day during which the surgery was performed, which could have potentially altered our findings. Future studies could benefit from incorporating these clinical factors to provide additional understanding of turnover time determinants.

We recommend efforts focusing on building a standardized environment for surgeons regardless of their background. This could lead to not only an equitable OR culture but also an overall increase in the institution’s efficiency and patient outcomes.

## Data Availability

Available upon request

## Notes

**Conflicts of interest/ Disclosures :** None

### Competing Interest Statement

The authors have declared no competing interest.

### Funding Statement

No funding

### Author Declarations

Our proposal was approved by our institution Aligning and Coordinating Quality Improvement, Research, and Evaluation (ACQUIRE) Committee, which approves all projects that are intended for local improvement of care. The ACQUIRE Committee is a dedicated ethics oversight body and approves all quality-improvement projects that don't require an IRB.

## References

1. Harders M, Malangoni MA, Weight S, Sidhu T. Improving operating room efficiency through process redesign. Surgery. 2006 Oct 1;140(4):509–16.

2. Cohen TN, Anger JT, Shamash K, Cohen KA, Nasseri Y, Francis SE, et al. Discovering the barriers to efficient robotic operating room turnover time: perceptions vs. reality. J Robotic Surg. 2020 Oct 1;14(5):717–24.

3. Gottschalk MB, Hinds RM, Muppavarapu RC, Brock K, Sapienza A, Paksima N, et al. Factors Affecting Hand Surgeon Operating Room Turnover Time. Hand (New York, N,Y). 2016 Dec 1;11(4):489–94.

4. Cendán JC, Good M. Interdisciplinary Work Flow Assessment and Redesign Decreases Operating Room Turnover Time and Allows for Additional Caseload. Archives of Surgery. 2006 Jan 1;141(1):65–9.

5. Zhong H, Zhou L, Liao S, Tang J, Yue L, Mo M, et al. Effects of a fixed nurse team in the orthopaedic surgery operating room on work efficiency and patient outcomes: a propensity score-matched historically controlled study. BMC Nurs. 2022 Sep 6;21(1):248.

6. Broeders JAJL, Draaisma WA, van Lanschot JJB, Broeders IAMJ, Gooszen HG. Impact of Surgeon Experience on 5-Year Outcome of Laparoscopic Nissen Fundoplication. Archives of Surgery. 2011 Mar 21;146(3):340–6.

