## Supplemental file for "Surgeon Factors and Their Association With Operating Room Turnover Time"

**Category of submission:** Research Letter

**Conflicts of interest/ Disclosures :** None

**Sources of funding**: None

**Data availability**: available upon request

**Corresponding author:**

Aditya Bagrodia, MD

Department of Urology

Moores UCSD Cancer Center

3855 Health Sciences Drive, Mail Code: 0987

La Jolla, CA 92093-0987, USA

Supplementary methods:

We conducted a retrospective study at the UC San Diego School of Medicine, a tertiary academic medical center. We analysed surgical case entries from January 2022 to July 2023, sourced from the hospital’s electronic health record system. Surgeons were categorized by gender, ethnicity, years of experience, training at UCSD, academic rank, and administrative roles.

We collected OR data from the hospital’s electronic health record system to obtain 65,160 case entries for consecutive case performed across all specialties by 307 surgeons affiliated to UCSD. It was calculated by summating six time components from the OR data: 1) Wrap up time after end of first case 2) Wheeling out to clean-up start after the first case 3) Clean-up after first case 4) Completion of clean-up to the start of second case set-up 5) Second case set-up and, 6) Wheeling in patient for the second case. Case entries where turnover time exceeded two standard deviations above the mean (127 minutes) were excluded to eliminate outliers caused due to extra-ordinary circumstances other than normal surgical workflow. Surgeons with less than 3 recorded case entries were also excluded to ensure a reliable representation of each surgeon’s performance. The exclusion process yielded a total of 12,820 case entries and 268 surgeons for the analysis. Data for surgeon-specific factors was collected through their official university profiles. The variables captured were ethnicity, gender, experience (recorded as a binary variable as greater or less than 10 years of experience), residency or fellowship training at UCSD, academic rank and administrative position. Administrative positions were defined if the surgeon served as the department chair or vice-chair, division chief or held a leadership role such as chief executive officer in the health administration system.

We utilized Mann-Whitney U test for binary variables and Kruskal-Wallis H test for variables with more than two categories. Multivariable linear regression was applied, adjusting for multiple comparisons using Holm’s correction. A p-value of less than 0.05 was considered statistically significant. All data analysis was performed using IBM SPSS version 29.

Our proposal was approved by our institution’s Aligning and Coordinating Quality Improvement, Research, and Evaluation (ACQUIRE) Committee, which approves all projects that are intended for local improvement of care.

Supplementary Table S1: Turnover time with subcomponents for surgeons stratified by administrative position and experience

|  | **Non-administrators** | **Administrators** | **p-value** |
| --- | --- | --- | --- |
| **Turnover time (minutes)**  **(Inter-quartile ranges)** | 35 (26,49) | 27 (20,38) | <0.001 |
| **Subcomponents (minutes)**  **(Inter-quartile ranges)** |  |  |  |
| **Wrap-up time after the first case** | 5 (2,8) | 5 (4,9) | <0.001 |
| **Wheeling out to clean-up start** | 2 (1,4) | 1 (1,2) | <0.001 |
| **Clean-up duration** | 7 (4,10) | 4 (3,6) | <0.001 |
| **Completion of clean-up to the start of second case setup** | 4 (2,7) | 5 (3,7) | <0.001 |
| **Second case setup** | 10 (6,16) | 5 (4,10) | <0.001 |
| **Wheeling in for the second case** | 7 (4,12) | 7 (4,12) | 0.071 |
|  | **Surgeons with less than 10 years’ experience** | **Surgeons with more than 10 years’ experience** |  |
| **Turnover time (minutes)**  **(Inter-quartile ranges)** | 37 (27,52) | 31 (23,44) | <0.001 |
| **Subcomponents (minutes)**  **(Inter-quartile ranges)** |  |  |  |
| **Wrap-up time after the first case** | 2(1,4) | 2 (1,3) | <0.001 |
| **Wheeling out to clean-up start** | 6 (4,10) | 4 (2,7) | <0.001 |
| **Clean-up duration** | 7 (5,10) | 6 (4,9) | <0.001 |
| **Completion of clean-up to the start of second case setup** | 4 (2,8) | 4 (2,8) | 0.49 |
| **Second case setup** | 11 (7,18) | 8 (5,15) | <0.001 |
| **Wheeling in for the second case** | 7 (4,13) | 7 (4,15) | <0.001 |
